# Cerebrospinal fluid Aβ42 and fractalkine are associated with Parkinson’s disease with freezing of gait

**DOI:** 10.1101/2020.12.16.20248342

**Authors:** J. M. Hatcher-Martin, J.L. McKay, B. Sommerfeld, J.C. Howell, F. C. Goldstein, W.T. Hu, S.A. Factor

**Author notes:** Address Correspondence to: Stewart A. Factor, DO, Professor and Director, Jean and Paul Amos Parkinson’s Disease and Movement Disorder Program, Vance Lanier Chair of Neurology Emory University School of Medicine 12 Executive Park Drive NE, Atlanta, GA 30329. These authors contributed equally to the manuscript (J. M. Hatcher-Martin, J.L. McKay, S.A. Factor). Funding: William N. and Bernice E. Bumpus Foundation, Curtis family Fund; Sartain Lanier Family Foundation; American Parkinson’s Disease Association; NIH K25 HD086276. Statistical analysis was completed by J. Lucas McKay, Ph.D., M.S.C.R. from Emory University. Study Funding: Curtis Family Fund, Sartain Lanier Family Foundation (SAF), Parkinson Foundation, NIH K25HD086276 (JLM), Parkinson’s Foundation Fellowship (SPP). Dr. Hatcher-Martin has the following disclosures. Honoraria: Acadia, Neurocrine, Parkinson’s Foundation. Dr. McKay has the following disclosures. Consulting fees: Biocircuit Technologies. Mr. Howell has nothing to disclose. Dr. Goldstein reports no disclosures. Dr. Hu has patent or patent-pending on CSF diagnosis of FTLD-TDP, CSF prognosis of SMA, and serological assays for COVID-19; licensed serological assays for COVID-19 to Sigma-Millipore; consulted for Biogen Inc., Fujirebio Diagnostics Inc., Apellis Pharmaceuticals, and AARP; and received research support from Fujirebio Diagnostics Inc. Dr. Factor has the following disclosures. Honoraria: Lundbeck, Teva, Sunovion, Biogen, Acadia, Neuroderm, Acorda, CereSpire. Grants: Ipsen, Medtronic, Boston Scientific, Teva, US World Meds, Sunovion Therapeutics, Vaccinex, Voyager, Jazz Pharmaceuticals, Lilly, CHDI Foundation, Michael J. Fox Foundation, Royalties: Demos, Blackwell Futura for textbooks, Uptodate. Authors roles. JM Hatcher-Martin: Co-first author, Research Project- Conception, Organization, Execution, Statistical analysis-review and critique, Manuscript: Review and critique, Arranged funding JL McKay: Co-first author, Research Project-Execution, Statistical analysis-Execution, Manuscript: Writing of the first draft, review and critique. B Sommerfeld: Research Project-Execution, Statistical analysis- no role, Manuscript: Review and Critique. JC Howell: Research Project-Execution, Statistical analysis- no role, Manuscript: Review and Critique. FC Goldstein: Research Project-Execution, Statistical analysis- Review and Critique, Manuscript: Review and Critique. WT Hu: Research Project-Execution, Statistical analysis- Review and Critique, Manuscript: Review and Critique. SA Factor: Research project: Conception, Organization, Execution, Statistical Analysis: Review and Critique; Manuscript: Writing of the first draft, Arranged funding.

## Abstract

**Objective:** To evaluate the association of Alzheimer’s disease-related and inflammation-related cerebrospinal fluid (CSF) markers with freezing of gait (FOG) in patients with Parkinson’s disease (PD).

**Methods:** The study population included well-characterized PD patients with FOG (PD-FOG), without FOG (PD-NoFOG) and healthy controls (HC). CSF was collected using standard protocols. Three Alzheimer’s disease-related markers and 10 inflammation-related markers were measured in a Luminex 200 platform. Differences in marker expression across groups were evaluated with multivariate linear models.

**Results:** CSF was collected from PD-FOG (N=12), PD-NoFOG (N=20) and HC (N=11) for analysis. Age was not significantly different between the three groups. Duration of PD was not significantly different between the two PD groups. After adjusting for covariates and multiple comparisons, the anti-inflammatory marker, fractalkine, was significantly decreased in the PD groups compared to HC (P=0.022), and further decreased in PD-FOG compared to PD-NoFOG or HC (P=0.032). The Alzheimer’s disease-related protein, Aβ42, was increased in PD-FOG compared to PD-NoFOG or HC (P=0.004). p-Tau_181_ was also decreased in both PD groups compared to HC (P=0.010).

**Conclusions:** We found high levels of Aβ42 in PD-FOG patients and cross-sectional data which supports an increase over time from early to advanced state. We also found low levels of fractalkine which might suggest anti-inflammatory effect. This is the first time an association between fractalkine and FOG has been shown. Whether these changes are specific to FOG requires further exploration.

## Introduction

Freezing of gait (FOG) is characterized by arrests of stepping when initiating gait, turning, and walking straight ahead and patients describe it as their feet being “glued” to the floor^1^. While it is a well-known feature of PD, it has been reported in other parkinsonian disorders as well^2^. FOG frequency in PD patients increases with disease duration and occurs in >60% of patients with >/=10 years of disease^3^. It is a gait symptom complex that has potentially grave consequences^1,4^ as it is unpredictable in character, a leading cause of falls with injury, and results in loss of independence and social isolation. Treatment options are limited^1,5^. Although FOG is considered to be a cardinal feature of PD, it appears to develop and/or progress independently of the other cardinal motor features^6^. It is associated with specific clinical risk factors (longer disease duration, psychotic symptoms, and absence of tremor), is associated with cognitive change and is thought to be caused by specific as yet unknown pathology^7,8 9^. The pathophysiology of FOG remains poorly understood. The literature shows great variability in findings related to physiological and imaging research, as well as motor and non-motor correlates and therapeutic response to various treatment modalities^8,10-12^ suggesting it possibly relates to a circuit change. There has been little in the way of biofluid research in FOG. One study of data from the Parkinson Progression Marker Initiative study (PPMI) showed that low CSF β-amyloid42 (Aβ42) levels in early PD predicted incident FOG within the first few years after diagnosis^13^ using measures from the Movement Disorder Society-Unified Parkinson’s disease Rating Scale (MDS-UPDRS) and this has been supported by increased neocortical deposition of β-amyloid in brain with imaging studies^14^. However, these data have been reported only for the first three years after disease onset. In this study we present initial results of CSF analysis in PD patients with and without FOG and aged-matched healthy controls. We also present results describing variation in CSF markers over a wide range of disease duration (onset–23 years).

## Results

### Participants

CSF samples were collected from PD-FOG (N=12), PD-NoFOG (N=20) and HC (N=11) for analysis. Age was not significantly different for the three groups and duration of PD was not significantly different for the PD-FOG and PD-NoFOG groups. The HC group included more females than either of the PD groups: 73% female in the HC group, 35% PD-NoFOG group and 8% PD-FOG group (**Table 1**).

**Table 1.** Demographic and clinical features of the study sample.

|  | HC | PD-NoFOG | PD-FOG |
| --- | --- | --- | --- |
| N | 11 | 20 | 12 |
| Age, y | 73.5 (9.9) | 71.1 (10.4) | 70.7 (8.3) |
| Sex<sup>a</sup> |  |  |  |
| Female | 8 (73%) | 7 (35%) | 1 (8%) |
| Male | 3 (27%) | 13 (65%) | 11 (92%) |
| PD duration, y |  | 9.7 (4.1)<sup>b</sup> | 11.5 (5.7) |

Data are presented as mean ± standard deviation or as N (%). <sup>a</sup>Significant difference between groups, Chi-squared test. <sup>b</sup>N=19.

### Variation in CSF marker expression with PD and FOG

Levels of CSF markers that varied across groups are summarized in **Table 2**. Expression of all CSF markers entered into analyses are summarized in **Table S2**. Initial univariate analyses identified four CSF markers associated with PD (p-Tau_181_, fractalkine, TGFα, MCP-1) and two CSF markers associated with PD-FOG (fractalkine, Aβ42). After adjustment for multiple comparisons and covariates (Figure 1), the anti-inflammatory marker, fractalkine, was significantly decreased in PD-NoFOG compared to HC (P=0.022, adjusted), and further decreased in PD-FOG compared to PD-NoFOG or HC (P=0.032, adjusted). The AD-related protein, Aβ42, was significantly increased in PD-FOG compared to PD-NoFOG or HC (P=0.004, adjusted). p-Tau_181_ was also significantly decreased in all PD compared to HC (P=0.010, adjusted) with no difference between those with and without FOG. No statistically significant effects of gender or PD duration were identified in models that were statistically significant for PD-NoFOG or PD-FOG, either before or after Holm adjustment.

**Table 2.** Average CSF marker levels that varied across groups.

|  | HC | PD-NoFOG | PD-FOG |
| --- | --- | --- | --- |
| N | 11 | 20 | 12 |
| Fractalkine<sup>a,b</sup> | 68.0 (11.7) | 59.1 (7.6) | 44.9 (17.3) |
| Aβ42<sup>b</sup> | 253.1 (92.2) | 200.3 (70.9)<sup>c</sup> | 354.7 (137.3) |
| p-Tau<sub>181</sub><sup>a</sup> | 24.7 (7.9) | 15.8 (9.2)<sup>c</sup> | 12.9 (5.0) |

Data are presented as mean ± standard deviation. <sup>a</sup>Significant difference between PD and HC, multivariate linear model with Bonferroni-Holm adjustment. <sup>b</sup>Significant difference between PD-FOG and other groups. <sup>c</sup>N=18.

**Figure 1.**
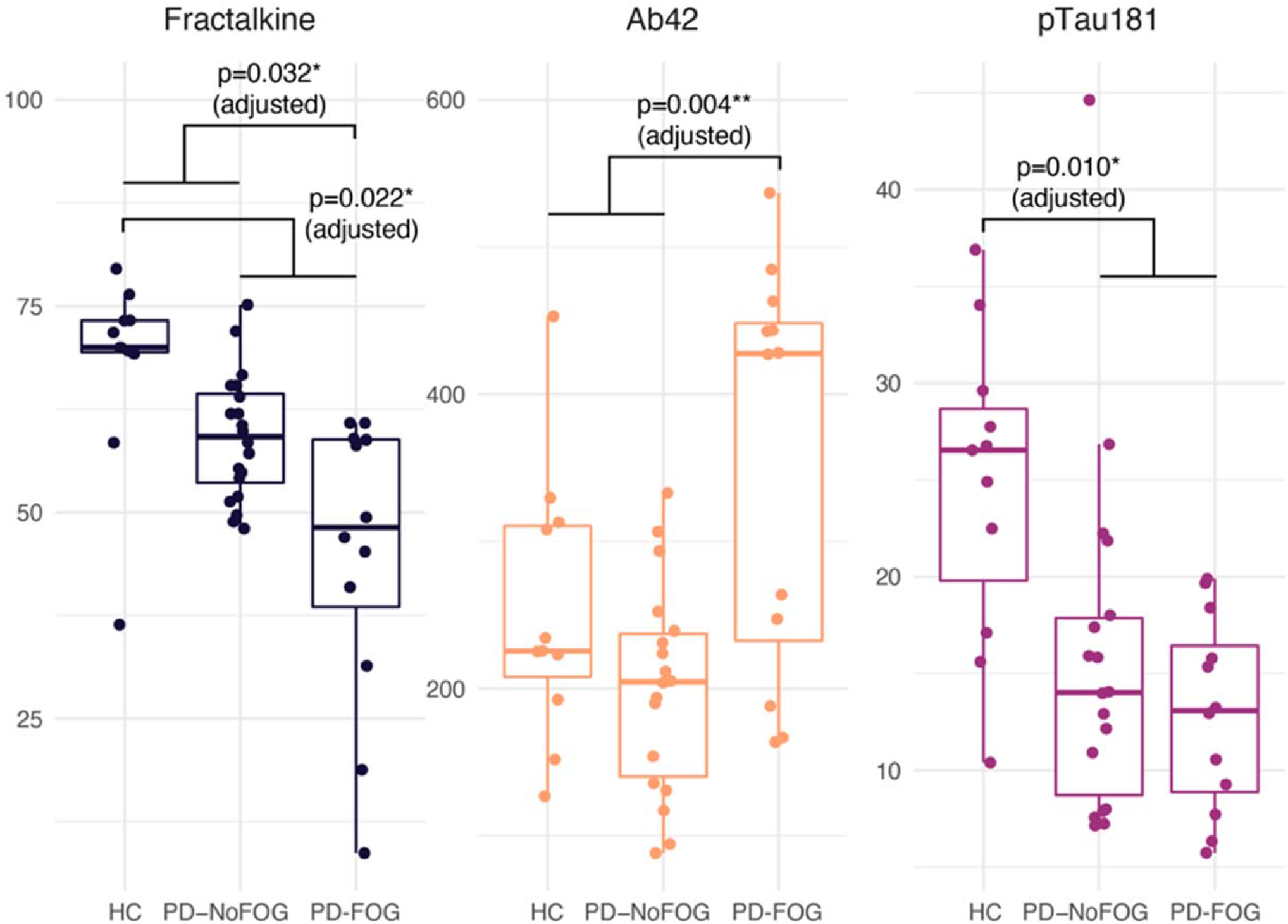
CSF marker levels. Box plots depicting the distribution of the CSF markers (fractalkine, Aβ42, and p-Tau_181_) across study groups. Boxes and horizontal lines depict ranges Q1-Q3 and median values of the expression of each marker, respectively. Abbreviations: HC, healthy control; PD-NoFOG, PD without FOG; PD-FOG, PD with FOG. P values reflect multivariate linear models controlled for covariates and multiple comparisons.

### Variation in CSF markers with disease duration

The association between each marker identified in initial multivariate analyses as varying across groups and PD duration is shown in **Figure 2**. There was a significant linear relationship (P=0.02) between Aβ42 expression and PD duration with significant (P=0.005) interaction between the PD-FOG and PD-NoFOG groups in multivariate models controlling for age and sex. Aβ42 increased with increasing duration in the PD-FOG group but decreased with increasing duration in the PD-NoFOG group. No other statistically significant associations were identified. Visual inspection of plots indicated some evidence of interaction between groups for fractalkine, in which the PD-FOG group decreased with increasing duration whereas the PD-NoFOG group increased somewhat, and no evidence of interaction for p-Tau_181_, in which both groups exhibited a decreasing relationship with PD duration.

**Figure 2.**
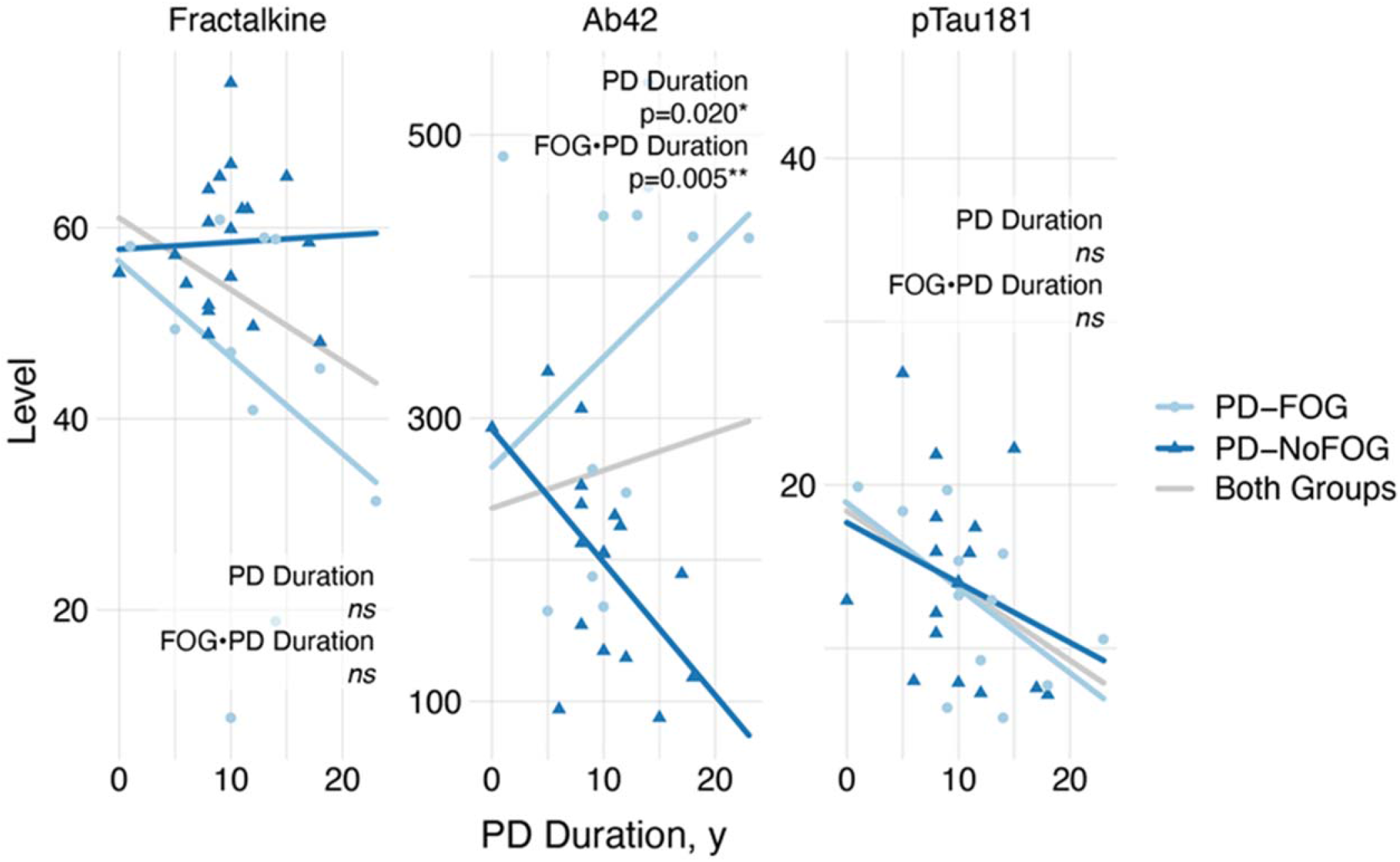
Association between CSF markers (Aβ42, fractalkine, and p-Tau_181_) and disease duration, stratified by presence of FOG. Light and dark blue lines represent separate linear regressions of marker level onto PD duration for PD-FOG and PD-NoFOG, respectively. Light gray lines represent linear regressions for both groups combined. P values reflect multivariate linear models with terms for age, sex, FOG, PD duration, and interaction between FOG and PD duration.

## Discussion

In this study we examined, in CSF, three AD-related markers and ten inflammation-related proteins in HC and in PD patients with and without FOG. Among those with PD and FOG – which was carefully characterized clinically and in our motion capture lab – multivariate models showed that Aβ42 was elevated compared to PD-NoFOG cases and to HC. We also found that the anti-inflammatory protein fractalkine was lower in PD-FOG vs PD-NoFOG and HC. While we identified reduced CSF p-tau among both PD groups vs. HC, which was consistent with multiple studies^15^, the changes in Aβ42 and fractalkine were unexpected and in the opposite direction than those seen in comparisons of AD and HC.

Aβ42 is a CSF marker that is low in AD^16^ and represents the first marker change^17^ that predicts the development of AD pathology. The low levels in AD may reflect increased accumulation in the brain, enhanced clearance, or some other yet unknown mechanisms^18^. Normally, there is a diurnal pattern of CSF amyloid, higher during wakefulness and lower during sleep. This diminishes with age and even more so in AD and relates to sleep disruption which is caused by Aβ42 aggregation seen in animal models^19^. In PD, much of the data on CSF markers comes from early-stage patients in PPMI. Aβ42 CSF levels in early PD (within 2 years of symptom onset) have been shown to be modestly lower than HC (≈9%, in baseline measures)^20,21,22^. At levels below a critical value, Aβ42 levels appear to be associated with earlier development and faster decline of cognitive function in PD patients^23^ especially in those with REM sleep behavior disorder^24^ or the *APOE* ε4 allele^25^. However, those with CSF Aβ42 levels closer to typical values, relationships between CSF Aβ42 and progression are less clear^26^. The association of low baseline CSF Aβ42 with early dementia in PD has been shown by other investigators as well^27^. One study suggested an association of low baseline levels to worse motor scores and, in particular, postural instability gait disorder (PIGD) sub score^22^ although such findings have not been consistently seen. The neuropathological correlates of lower CSF Aβ42 in PD also remain unclear. Whether those with low levels of CSF Aβ42 represent a subset of PD with coincident AD amyloid accumulation or altered amyloid metabolism in the brain remains to be seen, although previous *in vitro* animal and clinical cohort studies have suggested the increasingly important role of AD pathology in the development of PD Dementia^28^.

Our results suggest that the associations between lower CSF Aβ42 and incident FOG that hold in newly diagnosed PD patients^13^ may not generalize to older or more advanced patients. FOG is also associated with cognitive decline, particularly executive dysfunction^7-9^ which is associated with lower Aβ42. Our results in this study would appear to be in opposition to this finding, as we demonstrated higher CSF Aβ42 levels among the PD-FOG group. However, our CSF samples were taken in much later stages of disease than are currently available in studies like PPMI; the mean duration of the disease was 10.4 (±4.8) years and the longest duration in the sample was 23 years. Commensurate with their increased disease duration, these individuals were older than those in the most recent reports from PPMI (71.6 years vs. 61.4 years in PPMI)^22^. Additionally, among the FOG group here, there was no evidence of substantial executive dysfunction – the average MoCA score was 24.1 (results published separately^12^). This could suggest an increase in CSF levels of Aβ42 in the ensuing time frame, perhaps due to increased amyloid production – or decreased sequestration in the brain – particularly in the PD-FOG group. Our cross-sectional analysis of the relation between CSF Aβ42 and duration of disease in the PD-FOG and PD-NoFOG would also suggest that this increase is specific to those developing FOG, as we found a statistically significant interaction in the relationship between CSF Aβ42 and disease duration: Aβ42 increased with increasing duration in the PD-FOG group but decreased with increasing duration in the PD-NoFOG group. It has been shown in AD that longitudinal changes of CSF marker levels vary in distinct populations of subjects depending on underlying pathology^29^. There is limited data on longitudinal changes in levels over time in PD. Data from the PPMI study demonstrated an increase in Aβ42 over 6 and 12 months of follow-up in PD and HC groups which reached significance only at the one year mark compared to baseline (a 4% increase)^30^. This correlated with age in both groups and disease duration in the PD group. They did not examine subgroups such as those with FOG. Irwin et al more recently examined AD markers in PD and HC in a larger number of patients from the PPMI cohort and with a different assay from the prior assessment, this time followed up to three years. In longitudinal analysis they found that the PD cohort had a modestly greater decline in CSF Aβ42 (mean difference = −41.83 pg/mL; p = 0.03) and CSF p-tau (mean difference = −0.38 pg/mL; p = 0.03) at year 3 compared with HC^22^. These findings are consistent with our observation for PD-NoFOG. It is therefore possible that not only the baseline level but the pattern of change of Aβ42 may be predictive of the development of clinical outcomes, with increasing levels later in FOG. For example, in early PD, Kim et al. ^13^ recently showed moderately reduced (≈8%) CSF Aβ42 levels among patients who develop FOG within the first four years. In linear trends estimated from our data, a similar relationship is observed over the first ≈3 years, but after this point FOG is associated with increased, rather than decreased, CSF Aβ42 (Figure 2). Further longitudinal observations in PPMI subjects are needed to define the extent to which the prognostic performance of CSF markers varies with age or disease duration. It should be noted that high CSF Aβ42 levels is not unique to this PD-FOG cohort. It also appears in other scenarios including; slow wave sleep disruption ^31^, narcolepsy with cataplexy (especially those with normal CSF hypocretin-1 concentrations)^32^, particular gene polymorphisms in AD^33^, late-life depression^34, 35^, and traumatic brain injury^36^.

The existing evidence does not allow for definite conclusions regarding the role of fractalkine in PD pathophysiology, but our results suggest an anti-inflammatory effect with lower levels being associated with PD-FOG and with PD-NoFOG compared to HC. Fractalkine is a neuroimmune regulatory protein produced mainly by neurons and exists in native, membrane and soluble forms, each eliciting different cytokine responses from immune cells in the central and peripheral nervous systems. The soluble form, which is measured in this study, has a signaling function specifically through the G-protein-coupled CX3CR1 receptor that resides on microglia^37^. Fractalkine is known to have anti-inflammatory function under some circumstances as signaling contributes to suppressing microglial activation and maintaining the microglia surveillance phase^38^. As part of this function, it reduces the overproduction of proinflammatory molecules such as inducible nitric oxide synthase, interleukin (IL)-1β, (TNFα), and IL-6 generated by microglia^37,38^. However, in rodent toxin models it has been shown that the exact effects greatly depend on the isoform type (soluble or membrane-bound), animal model (mice or rats, toxin- or proteinopathy-induced), route of toxin administration, time course, specific brain region (striatum, substantia nigra), and cell type^38^. The same is true with regard to α-synuclein models, inflammatory response type depends on the form of α-synuclein (overexpressed wild-type or A53T-mutated form), the specific isoform of fractalkine and the experimental protocol^38^. Whenever neuroprotective, the soluble, and not the membrane-bound form of fractalkine seems to be responsible for its beneficial role^38^. Similar variation in results in AD models has been reported ^39^. In one study, in transgenic mouse models of AD (amyloid precursor protein/- presenilin1 and CX3CR1−/−), fractalkine brought about a decrease in amyloid burden^40^.

One previous CSF study in PD found no difference in fractalkine levels between HC and PD, but it is not clear if participants with FOG were included, which may explain the discrepancy between these and earlier results^41^. The fractalkine/Aβ42 ratio was weakly correlated with PD severity in cross-sectional and longitudinal PD samples. The reason for this may include assay differences, pre-analytical processing, and freeze-thawing effects. In another study, exosomal levels of fractalkine mRNAs were shown to be lower in the CSF of PD patients compared to HC^42^.

The main limitation of this study was the small sample size. Nevertheless, this was the first attempt to examine CSF markers in advanced PD patients with FOG. Another limitation was the cross-sectional nature. Longitudinal studies to examine the trajectory of change of fractalkine and Aβ42 markers will be important.

In conclusion, we examined AD and inflammatory-related CSF markers in advanced PD patients with and without FOG. We found high levels of Aβ42 in PD-FOG, and cross-sectional data which supports an increase over time from early to advanced state in the PD-FOG groups specifically. Longitudinal studies are needed to confirm this. Such results support a previously reported role of Aβ42 in the development of FOG. We also found low levels of fractalkine which might suggest an anti-inflammatory effect. This is the first time an association between fractalkine and FOG has been shown. Whether these changes are specific to FOG or relate to the cognitive change often associated with FOG or simply progression of disease requires further exploration.

## Methods

### Patients

The study population included PD patients with FOG (PD-FOG) and without FOG (PD-NoFOG) and healthy controls (HC). Parkinson’s participants and HC were recruited from the Emory Movement Disorders and Cognitive Neurology Clinics. Subject recruitment and assessments for those with PD-FOG and PD-NoFOG have been previously described^12^. Inclusion criteria for all PD patients were: Age≥18 years; PD diagnosis according to United Kingdom Brain Bank criteria^43^; Hoehn & Yahr stage I-IV in the OFF state; Demonstrated response to levodopa; Able to sign a consent document and willing to participate in all aspects of the study. Additional inclusion criteria for PD-FOG participants were: FOG noted in medical history and confirmed visually by examiner. Exclusion criteria were: Atypical parkinsonism; other neurological or orthopedic disorders interfering with gait; dementia or other medical problems precluding completion of study protocol. To demonstrate FOG patients underwent a rigorous assessment in the practically defined OFF state^44^. They then received a levodopa challenge and were assessed in the same manner in the full ON state^12^. Outcomes were quantified in clinic using the MDS-UPDRS part 3 scale and in a motion capture laboratory to be assured of the presence of FOG. Additional PD-NoFOG patients and HC were age matched and selected from the Emory Goizueta Alzheimer Disease Research Center^45^.

### Standard Protocol Approvals, Registrations, and Patient Consents

This study was approved by the institutional review board of Emory University. All subjects provided written informed consent.

### CSF analysis

CSF (20 mL) was collected using protocols modified from the Alzheimer’s Disease Neuroimaging Initiative (ADNI)^46^ using 24G Sprotte atraumatic needles and syringe between 8AM and noon without overnight fasting and transferred into two 15 mL polypropylene tubes. CSF was immediately aliquoted (500 μL), labeled, and frozen (−80 °C) until analysis (Fujirebio, Ghent, Belgium)^45^.

Established CSF Alzheimer’s disease (AD) markers (Aβ42, total tau [t-Tau], and tau phosphorylated at threonine 181 [p-Tau_181_]) were measured using AlzBio3 assays (Fujirebio Diagnostics Inc., Malvern, PA) in a Luminex 200 platform ^47^. In addition, ten inflammation-related proteins were selected for their preferential association with innate immunity or different immune cell populations: proinflammation cytokines included tumor necrosis alpha (TNFα), interleukin 7 (IL-7), interleukin 8 (IL-8), transforming growth factor alpha (TGFα), interferon gamma-induced protein 10 (IP-10), monocyte chemoattractant protein 1 (MCP-1); anti-inflammatory proteins included macrophage-derived chemokine (MDC), interleukin 9 (IL-9), interleukin 10 (IL-10) and fractalkine. All these proteins were measured in a Luminex 200 platform using the Merck-Milliplex MAP Human Cytokine Panel (HCYTOMAG-60K, Merck-Millipore, Burlington, MA) following the manufacturer’s protocol. All operators were blinded to the diagnosis^45^. In our laboratory, we achieve average intermediate precision (over experiments performed over 9 days) of 9.4% for TNF-α, 12.9% for MDC, 14.7% for IL-7, 4.8% for IP-10, 12.0% for IL-10, 9.2% for IL-9, and 7.6% for IL-8.

### Statistical Analysis

Differences in demographic and clinical variables across groups were assessed with tests of central tendency (chi-square, ANOVA). Levels of 13 putative CSF markers (**Table S1**) were measured using separate assays and compared across participants with and without PD and with and without FOG using linear models including terms for sex and PD duration. PD duration in years was centered about 0 prior to entry in linear models such that HC were coded with a value of 0, PD patients with above average PD duration were coded with a positive number, and PD patients with below average PD duration were coded with a negative number. The value 0 (representing the average duration) was imputed for one participant with PD for whom this data point was unavailable. P values for factors representing presence of PD and presence of FOG were calculated and adjusted to control Type I error with a Bonferroni–Holm procedure.

In order to assess variation of marker expression with disease duration among individuals with PD, markers that were identified in univariate analyses as varying across groups were entered into multivariate regression analyses with terms for PD duration, FOG, age, and sex. Interaction terms were included to examine whether different relationships between marker expression and disease duration were apparent in PD-NoFOG and PD-FOG. All statistical analyses were performed in R software at alpha=0.05.

Data Availability Statement: Data not available in this manuscript will be shared at the request of other qualified investigators for purposes of replicating procedures and results.

## Data Availability

Data not available in this manuscript will be shared at the request of other qualified investigators for purposes of replicating procedures and results.

